# GC801-enabled AAV gene therapy in a neonate with Pompe disease

**DOI:** 10.64898/2026.09.23.26363499

**Authors:** Jia Chen, Wenhao Ma, Hongbo Qi, Xiuwei Ma, Yanmei He, Liu He, Yan Ke, Lu Zhuang, Xinyang Jiang, Jianhua Wang, Juan Xu, Yan Xia, Jianfang Zhou, Yue Wang, Xiaoyan Dong, Wu Xiaobing, Zhichun Feng

## Abstract

We report a neonate with IOPD who had high pre-existing anti-AAV9 antibody titers, which precluded eligibility for AAV9-mediated gene therapy (GC301). Preconditioning with a single dose of the IgG-degrading enzyme GC801 (0.2 mg/kg) safely reduced serum IgG from 6.25 g/L to a nadir of 0.72 g/L within 5 days. This pharmacological window facilitated successful GC301 vector transduction, driving a robust increase in GAA activity to 19.38 mmol/L/16h by day 84. Crucially, no adverse events beyond the underlying IOPD were observed. Our findings support the potential utility of GC801 as an agent to overcome pre-existing humoral barriers in neonatal gene therapy candidates.

---

Adeno-associated virus (AAV) vectors are a cornerstone of modern gene therapy,^1,2^ yet their efficacy is frequently undermined by pre-existing neutralizing antibodies (NAbs) against capsid proteins.^3,4^ Present in approximately one-third of the population, these NAbs trigger rapid vector clearance, creating a major translational bottleneck by excluding a large subset of patients from treatment.^3-5^ Existing mitigation strategies—such as plasmapheresis or FcRn antagonism—suffer from limitations including cost, off-target immunosuppression, and slow onset.^5,6^ The *Streptococcus pyogenes*–derived enzyme IdeS offers a compelling alternative by specifically cleaving the IgG hinge region, leading to immediate and irreversible IgG depletion.^3^ Although its recombinant form (imlifidase) is clinically approved for desensitization in adult kidney transplantation, and has shown promise against AAV8/AAV3B NAbs, its use in pediatric patients remains unexplored.^7^

This challenge is particularly acute in pediatric gene therapy, where early intervention is essential but often impeded by maternally derived antibodies.^8^ Here, we describe the first-in-neonate use of GC801, an engineered IdeS variant developed by Beijing Genecradle Therapeutics. Administered prior to AAV9 gene therapy for infantile-onset Pompe disease (IOPD), GC801 effectively mitigated pre-existing anti-AAV9 NAbs.

## Result

### Subject Characteristics

The diagnosis of IOPD was confirmed through integrated prenatal and postnatal evaluations: amniocentesis with whole exome sequencing identified compound heterozygous pathogenic GAA variants (c.1848C>G and c.2237G>A), while prenatal ultrasonography revealed progressive biventricular hypertrophy; postnatally, profoundly reduced plasma GAA activity (0.52–0.56 mmol/L/16h) and severe multisystemic involvement—including hypertrophic cardiomyopathy, sinus bradycardia, pulmonary hypertension, and renal and hepatic dysfunction—were observed (Table 1 and Figure 1A). Baseline biochemistry underscored the disease severity, with markedly elevated myocardial injury markers (NT-proBNP: 12,328 pg/mL; hs-Troponin: 19.91 pg/mL), significant skeletal muscle damage (CK: 2473 U/L; LDH: 1136 U/L), and metabolic disturbances (creatinine: 112.30 μmol/L; bilirubin: 80.60 μmol/L). Although AAV9-mediated GAA gene therapy was indicated based on our institutional experience, screening revealed high titers of anti-AAV9 binding (1:4174.2) and neutralizing antibodies (1:672.6), signifying a substantial pre-existing humoral barrier. Consequently, the patient received GC801 infusion to deplete systemic IgG, thereby establishing optimal transduction conditions for subsequent GC301 gene therapy.

**Table 1.** Clinical Characteristics, Medical History, and Safety of IdeS Treatment.

|  |  |
| --- | --- |
| Period□ | Details□ |
| Date of Birth□ | Blurred for ethical reasons |
| Sex | Blurred for ethical reasons |
| *Prenatal Period*□ |  |
| Antenatal Ultrasound | Progressive biventricular hypertrophy, worsening tricuspid regurgitation (mild to moderate), a mildly increased cardiothoracic |
| Pathogenic GAA Variants | c.1848C>G (p.Asp616Glu) and c.2237G>A (p.Trp746*) |
| Obstetric History | Vaginal birth, GA 37+5 weeks, Apgar 10/10 |
| Postnatal Vital Signs (0–2 h)□ | HR 90–110 bpm; SpO<sub>2</sub> 90%–95% |
| *Pre-Gene Therapy Admission*□ |  |
| Diagnoses□ | 1. Glycogen storage disease type II; 2. Cardiac hypertrophy; 3. Sinus bradycardia; 4. Prolonged QT interval; 5. Abnormal cardiac enzyme profile; 6. Patent ductus arteriosus; 7. Neonatal pulmonary hypertension; 8. Renal dysfunction; and 9. Hepatic dysfunction |
| *Laboratory Baseline (Pre-treatment)*□ |  |
| Plasma GAA activity, mmol/L/16h | 0.52 ~ 0.56 |
| NT-proBNP, pg/mL | 12,328 (reference range: 0–100) |
| sST2, ng/mL | 164.64 (35–50) |
| Myoglobin, ng/mL | 710.90 (<90) |
| H-FABP, ng/mL | >120 (<6) |
| hs-Troponin, pg/mL | 19.91 (<14) |
| CK, U/L | 2473 (<200) |
| CK-MB, U/L / ng/mL | 119.3 (<25) |
| LDH, U/L | 1136 (120 - 250) |
| α-HBDH, U/L | 757 (72 - 182) |
| AST, U/L | 257.18 (21 - 80) |
| Total Bilirubin, μmol/L | 80.60 (<21) |
| Serum Creatinine, μmol/L | 112.30 (13 – 33) |
| *Three-Month Safety Follow-Up After IdeS Desensitization and GAA Gene Therapy*□ |  |
| Adverse Events (AEs)□ | No AEs beyond those attributable to the underlying IOPD were observed (assessed per Common Terminology Criteria for Adverse Events [CTCAE] version 5.0) |
| Abbreviations: GAA, acid alpha-glucosidase; HR, heart rate; SpO<sub>2</sub>, saturation of peripheral oxygen; GA, gestational age; NT-proBNP, N-terminal pro-B-type natriuretic peptide; sST2, soluble suppression of tumorigenicity 2; H-FABP, heart-type fatty acid binding protein; hs-Troponin, high-sensitivity cardiac troponin; CK, creatine kinase; CK-MB, creatine kinase-MB; LDH, lactate dehydrogenase; α-HBDH, α-hydroxybutyrate dehydrogenase; AST, aspartate aminotransferase. |  |

**Figure 1.**
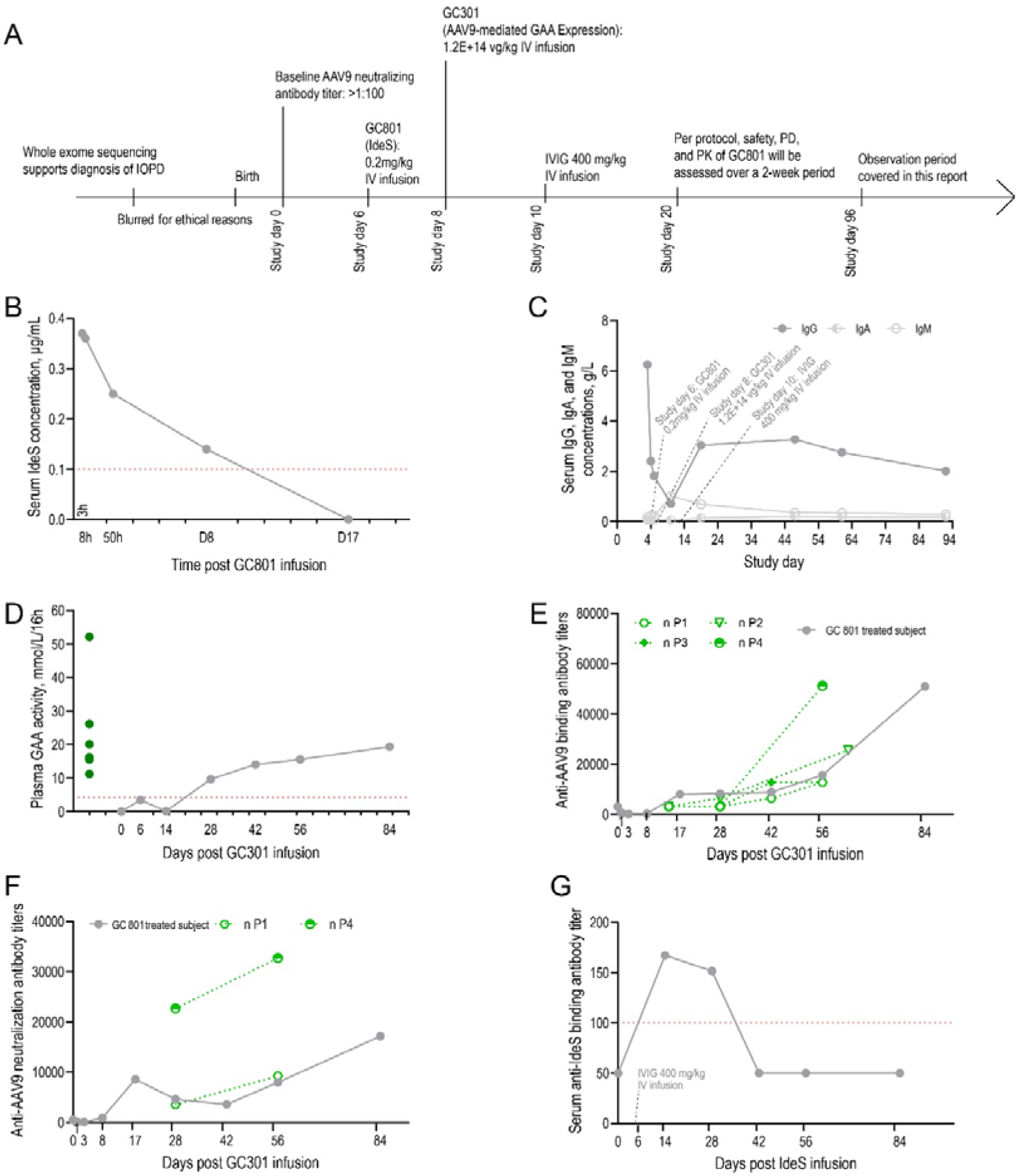
Pre-emptive reduction of pre-existing anti-AAV9 antibodies with GC801 enables effective GAA delivery and associated pharmacokinetic and pharmacodynamic profiles. (A) Timeline of patient history, dosing schedule for GC801, GC301, and IVIG, and the temporal framework for safety and efficacy monitoring. (B) Pharmacokinetic profile of GC801 (IdeS). The red dashed line indicates the lower limit of quantification. (C) Kinetic changes in serum immunoglobulin levels (IgG, IgM, and IgA) following GC801 administration. (D) Temporal dynamics of plasma acid α-glucosidase (GAA) activity following GC301 infusion. Green dots denote reference GAA levels in healthy adults. (E) Longitudinal profiles of anti-AAV9 binding antibody titers following GC301 infusion. Green dashed lines represent corresponding titers from four historical patients receiving GC301 gene therapy. (F) Longitudinal profiles of anti-AAV9 neutralizing antibody titers following GC301 infusion. Green dashed lines represent corresponding titers from two historical patients receiving GC301 gene therapy. (G) Temporal kinetics of serum anti-IdeS antibody development post-GC801 infusion. Abbreviations: GA, gestational age; PD, pharmacodynamics; PK, pharmacokinetics; IVIG, intravenous immunoglobulin; h, hour; D, day.

### Safety Profile

GC801 was administered as a single intravenous infusion a dose of 0.2 mg/kg prior to GC301 gene therapy. Throughout the 2-week observation period (and extending into the 90-day follow-up), GC801 exhibited a favorable safety profile. No AEs attributable to GC801 were observed (Table 1).

### Efficacy Profile

GC801 concentrations peaked at 0.37 µg/mL 3 hours post-infusion, remained above 0.25 µg/mL at 50 hours, and declined to the lower limit of detection (LoD) by approximately day 14. This pharmacokinetic profile served as a key scientific rationale for establishing the 14-day safety and efficacy observation period (Figure 1B).

Serum IgG was 6.25 g/L at baseline. Following GC801 infusion, levels decreased to 2.41 g/L and 1.82 g/L at 24 and 48 hours, respectively (coinciding with GC301 administration at ∼48 hours). By day 5 post-GC801, IgG further declined to 0.72 g/L. To mitigate complications of hypogammaglobulinemia, the infant received prophylactic IVIG (400 mg/kg) on day 7. This intervention raised IgG to 3.04 g/L by day 14, with concentrations subsequently stabilizing between 2.02 and 3.28 g/L for the remainder of the observation period (Figure 1C). Concurrent measurements of IgA and IgM demonstrated that GC801 selectively depleted IgG without affecting other immunoglobulin isotypes (Figure 1C). GAA activity increased from 9.67 mmol/L/16 h on day 28 to 19.38 mmol/L/16 h on day 84 post-GC301 infusion. This elevation not only restored GAA levels to within the reference range of healthy controls (11.21–52.14; median 18.17) but also suggested that GC801-mediated preemptive immunodepletion facilitates transgene-derived GAA expression (Figure 1D).

### Post-treatment antibody responses

The anti-AAV9 IgG responses exhibited an upward trend beginning 8 days post-GC301 infusion and were interpreted in two distinct phases: an initial modest rise (days 8–42), attributable to prophylactic IVIG infusion, followed by a subsequent steep rise (Figures 1E and 1F). Notably, data from four previously treated IOPD patients (aged 2.2–5.5 months) showed a similar kinetic profile. This indicates that GC801 conditioning did not substantially alter de novo antibody responses. Finally, serum anti-IdeS binding antibody analysis revealed a biphasic kinetic profile characterized by a transient elevation attributable to prophylactic IVIG infusion, flanked by periods during which levels remained below the LoD (Figure 1G).

## Discussion

This report describes the first-in-neonate application of GC801, an engineered IdeS protease, to overcome pre-existing humoral immunity against AAV9 capsids. Although IOPD provided the clinical context, the primary objective was to evaluate the safety and efficacy of GC801 as a desensitization platform for neonatal gene therapy; consequently, disease-specific outcomes (e.g., motor milestones or survival) fall outside the scope of this analysis.

GC801 demonstrated a highly favorable safety profile. A single intravenous dose of 0.2 mg/kg was not associated with any AEs attributable to the enzyme. Pharmacodynamically, GC801 induced rapid and profound IgG cleavage, reducing serum IgG from 6.25 g/L to a nadir of 0.72 g/L by day 5, thereby establishing a pharmacological window permissive for AAV9-mediated gene transfer. Crucially, this depletion was isotype-specific; IgA and IgM levels remained stable, preserving components of the neonatal immune repertoire.^9^ Transient hypogammaglobulinemia was effectively mitigated by prophylactic IVIG administered on day 7. These findings establish the feasibility of GC801 preconditioning in early neonates—a population previously unexplored for IdeS-based therapies.^7,10^

The increase in plasma GAA to 19.38 mmol/L/16h by day 84 (within the reference range of healthy controls) serves as a direct pharmacodynamic readout confirming successful AAV9-mediated transgene expression. This biochemical correction is mechanistically linked to GC801’s action: by cleaving circulating IgG, including pre-existing anti-AAV9 antibodies, GC801 prevented immediate vector neutralization and clearance, thereby enabling somatic cells transduction and sustained GAA production.^4^

Analysis of the immune response further delineates GC801’s immunologic footprint. Serum GC801 concentrations peaked at 0.37 µg/mL at 3 hours post-infusion and declined below the LoD by day 14, correlating with the transient nature of IgG depletion. Anti-IdeS antibody responses remained largely undetectable throughout the observation period; a transient, low-level elevation temporally aligned with IVIG administration likely reflected assay cross-reactivity with infused donor antibodies rather than de novo anti-drug antibodies.^11^ Importantly, the kinetics of de novo anti-AAV9 antibody development post-GC301 mirrored those in historical controls receiving GC301 without desensitization. This similarity indicates that GC801 preconditioning does not perturb subsequent humoral responses to the AAV9 capsid.^12^ In conclusion, GC801 represents a targeted strategy to mitigate pre-existing anti-AAV immunity in neonatal gene therapy candidates. Its favorable safety profile, rapid onset, and IgG specificity address key limitations of existing desensitization approaches, such as plasmapheresis or broad immunosuppression. Future studies are warranted to define optimal dosing windows and applicability across diverse AAV serotypes.

## Data Availability

ll data produced in the present study are available upon reasonable request to the authors

https://www.chictr.org.cn/showproj.html?proj=315073

## METHODS

This single-arm, open-label, single-center investigator-initiated trial (Chinese Clinical Trial Registry identifier: ChiCTR2600120964 and ChiCTR2600120429) was designed to evaluate the safety and preliminary efficacy of GC801 for the depletion of pre-existing anti–AAV9 antibodies in pediatric patients who were otherwise ineligible for AAV-mediated gene therapy. The study protocol was approved by the Ethics Committee of the Chinese PLA General Hospital (approval number S2025-100-02 and S2026-036-02) and conducted in accordance with the Declaration of Helsinki. Written informed consent was obtained from the legal guardians of participant prior to enrollment. We planned to enroll three patients aged 0 to 8 years; the present report describes the sentinel case—a neonate with IOPD and pre-existing anti-AAV9 immunity. Eligible participants were required to have serum anti-AAV9 binding antibody titers ranging from 1:1000 to 1:10,000 (inclusive), with IgG levels within the normal age-adjusted range at screening. Key exclusion criteria included a history or family history of primary or secondary immunodeficiency, hypersensitivity to streptococcal or protease-derived agents, severe allergic reactions or anaphylaxis, significant intercurrent illness within 4 weeks before enrollment, vaccination within 2 weeks before enrollment, or any other condition deemed unsuitable by the investigator. Descriptive statistics were used to summarize all safety and efficacy data.

GC801, a cysteine protease variant derived from *Streptococcus pyogenes* IdeS and developed by Beijing Genecradle Therapeutics,^1^ was administered as a single intravenous infusion at a dose of 0.2 mg/kg. This agent mediates the rapid, transient cleavage of human IgG into F(ab’)□ and Fc fragments, thereby disrupting its structural integrity and effector functions.^1,2^ Pharmacodynamic assessment was performed after GC801 administration; if total serum IgG decreased to less than 50% of baseline values, the patient proceeded to receive the GC301. GC301 is an AAV9 vector encoding codon-optimized human GAA complementary DNA and was administered intravenously at a dose of 1.2 × 10^1^□ vg/kg approximately 48 hours following GC801 infusion. Given the anticipated nadir in serum IgG following IdeS-mediated cleavage, with recovery expected to begin 48 to 72 hours post-treatment, intravenous immunoglobulin (IVIG) at a dose of 400 mg/kg was administered on day 7 to mitigate the risk of hypogammaglobulinemia-related complications. Patients were monitored intensively throughout the peri-infusion period according to the study protocol.

Safety evaluations included continuous vital sign monitoring, serial laboratory assessments, and systematic adverse event recording.^3,4^ Efficacy assessments comprised serial measurements of total serum IgG, anti-AAV9 binding antibodies, and neutralizing antibody titers at baseline and on days 1, 2, 7, and 14 post-GC801 administration. Serum concentrations of GC801/IdeS were quantified at 3, 8, and 50 hours and on days 8 and 17. Plasma GAA enzyme activity was measured at baseline (day 0) and on days 6, 14, 28, and 60 following GC301 infusion. Detailed assay methodologies have been described previously.^1,3,4^

GC801 and GC301 were supplied by Genecradle Therapeutics.^1,3,4^ Study design was collaborative between five academic investigators and four representatives from Genecradle Therapeutics. Data collection and analysis were performed jointly by the investigators and company representatives; however, Genecradle Therapeutics had no further role in data interpretation or manuscript preparation. All authors had full access to the study data and vouch for the accuracy and completeness of the data and the fidelity of the study to the protocol.

## Acknowledgements

We thank all coordinators involved in this study, as well as the patient and her family.

## Author contributions

X.D., X.W., and Z.F. conceptualized the study. X.D., X.W., and W.M. designed and produced the gene therapy product GC801. J.C. and H.Q. served as the lead investigators for this clinical trial. X.M., Y.H., L.H., Y.K., L.Z., X.J., J.W., J.X., Y.X., and J.Z. coordinated the clinical study execution, data monitoring, safety monitoring, and laboratory assessments. Y.W. drafted the original manuscript. All authors contributed to data analysis, reviewed and edited the manuscript, and approved its submission.

